# Transspinal stimulation preceding assisted step training reorganizes neuronal excitability and function of inhibitory networks in spinal cord injury: A randomized controlled trial

**DOI:** 10.1101/2025.06.11.25329338

**Authors:** Abdullah M. Sayed Ahmad, Morad Zaaya, Noam Y. Harel, Maria Knikou

**Author notes:** Corresponding Author Information: Maria Knikou, PT, MBA, PhD, Professor of Neurosciences & Rehabilitation, Deputy Chair of Research and Faculty Development, Director of the Klab4Recovery Research Program, PhD Biology, Neurosciences/Graduate Center of CUNY, DPT Department/College of Staten Island, 2800 Victory Blvd, 5N-207, NY 10314.

## Abstract

**Introduction:** In this pilot randomized sham-controlled clinical trial, we characterized the spinal neuronal and network excitability in human spinal cord injury (SCI) when transspinal stimulation preceded locomotor training within the same session.

**Methods:** Fourteen participants with chronic SCI received an average of 40 sessions with 30 Hz transspinal stimulation delivered for 30 minutes during standing (active: n= 4; sham: n= 5) or supine (active: n= 5) followed by 30-minutes of robotic assisted step training. Before and 1-2 days after completion of all training sessions, we assessed the soleus H-reflex homosynaptic depression and soleus H-reflex recruitment curve, and the amount of reciprocal and presynaptic inhibition following conditioning stimulation of the antagonistic common peroneal nerve.

**Results:** Transspinal stimulation administered before locomotor training increased the amount of homosynaptic depression in the active-standing and active-supine groups, while presynaptic inhibition exerted on Ia afferent terminals increased in all study groups. Reciprocal Ia inhibition improved in the sham-standing and active-supine groups while in all groups the excitability threshold of muscle group Ia afferents was decreased in all groups.

**Conclusion:** This study demonstrated that transspinal stimulation preceding locomotor training partially restores natural spinal inhibition and produces network reorganization in chronic SCI. Noninvasive transspinal stimulation can increase the benefits of locomotor training, bringing spinal neuronal networks to a more functional state in chronic SCI.

## 1 Introduction

After spinal cord injury (SCI), synergistic neuromodulation therapies such as the combination of spinal cord stimulation with activity-dependent protocols like locomotor training are in great need to maximize recovery of somatic and non-somatic body functions. Spinal circuitry along with spinal integration of corticospinal drive are greatly impaired after SCI (Knikou 2007; Arvanian et al. 2009; Barthelemy et al. 2010; Tansey et al. 2012). The pathological dysfunction of neuronal mechanisms and altered excitation state of motoneurons after SCI are well documented. For example, low-frequency dependent soleus H-reflex depression (also known as homosynaptic, rate-dependent or post-activation depression), which is a natural form of inhibition in response to repetitive excitation of group Ia afferents, is significantly reduced or abolished after SCI and linked to spasticity (Grey et al. 2008; Thompson et al. 1998), prevents physiological modulation of muscle contraction following repetitive afferent discharges. Homosynaptic depression acts on synapses between Ia afferent terminals and motoneurons mediated by a decrease in the quanta of presynaptic neurotransmitter release (Crone and Nielsen 1989; Hultborn et al. 1996). In a similar manner, altered presynaptic inhibition contributes partly to reflex hyperreflexia after SCI (Calancie et al. 1993). Presynaptic inhibition is a selective, powerful spinal mechanism that filters sensory afferent feedback to prevent overexcitation of spinal cord motor neurons and thus plays a pivotal role in the regulation of movement (Côté et al. 2018; Rudomin 2009; Nielsen 2016). Reduced natural reciprocal Ia inhibition exerted between antagonistic muscles during voluntary movements represents another detrimental maladaptive reorganization of spinal neuronal networks after SCI resulting in hyperreflexia, spasticity, co-contractions, and poor recovery of locomotion (Crone et al. 1994; Crone and Nielsen 1994; Morita et al. 2001).

Tonic noninvasive thoracolumbar transspinal (or transcutaneous spinal cord) stimulation may strengthen neural connectivity and thereby target pathological excitability in a manner similar to that described for epidural stimulation (Hofstoetter et al. 2017; Tajali et al. 2024). Transspinal stimulation at frequencies up to 15 Hz is used mostly for recovery of standing, while intermediate frequencies (25 - 120 Hz) are employed for recovery of stepping after SCI (Minassian et al. 2016; Rejc et al. 2017; Sayenko et al. 2019; Zaaya et al. 2021). Importantly, tonic transspinal stimulation is delivered at intensities and rates that modulate motoneuron membrane polarity and excitability without necessarily producing an action potential with each pulse. Significant effects on spinal locomotor pathways have been reported with 30 Hz transspinal stimulation in non-injured subjects (Skiadopoulos and Knikou 2024). Further, in several cases of complete or incomplete SCI, one (1) session of transspinal stimulation decreased spasticity, hyperreflexia, and ankle clonus, and increased leg muscle activity during stepping (Hofstoetter et al. 2014, 2015; Minassian and Hofstoetter 2016). Multi-session transspinal research studies are scarce, with the exemption of transspinal stimulation as single pulses at 0.2 Hz at intensities producing intermittent depolarization of alpha motoneurons (Knikou and Murray 2019; Murray and Knikou 2019). This protocol decreased soleus H-reflex excitability, upregulated homosynaptic inhibition, decreased spasticity, and increased the net motor output of motoneurons over multiple segments in people with SCI (Knikou and Murray 2019; Murray and Knikou 2019).

Alghouth insufficent at restoring full functional recovery, locomotor training alone also helps to restore spinal inhibition and decrease hyperreflexia in SCI (Knikou and Mummidisetty 2014; Knikou et al. 2015a; Smith et al. 2015; Smith and Knikou 2016). These changes were most likely due to plasticity of the glycinergic spinal inhibitory system that can occur independently of supraspinal influences (Sadlaoud et al. 2010). This is supported by animal studies that have shown that locomotor training normalizes the proportion of excitatory and inhibitory synaptic inputs to spinal motoneurons (Ichiyama et al. 2011; Rossignol and Frigon 2011), improves synaptic inputs from Ia afferents (Petruska et al. 2007) and alters neurotransmitter concentration (Ilha et al. 2011). Based on these parallels in effects on spinal circuitry inhibitory mechanisms, we suggest that transspinal stimulation may use similar mechanisms to that of exercise-dependent plasticity and thus act synergistically with locomotor training.

Many questions remain regarding how to combine transspinal stimulation with locomotor training. Should stimulation occur *during* training, or is it better delivered *before* training as a ‘primer’? Many multimodal studies have coupled locomotor training with central nervous system stimulation, *during* assisted stepping with substantial improvements in walking speed, spasticity, neuronal excitability, and motor output (Kumru et al. 2016; Raithatha et al. 2016; Pulverenti et al. 2021, 2022a, 2022b). A smaller number of studies have suggested that stimulating the nervous system *before* physical activity may prime the system to respond more effectively to task-specific exercise (Avenanti et al. 2012; Ackerley et al. 2016; Jo et al. 2023; Lotze et al. 2017; Estes et al. 2017). For example, Jo and colleagues showed that an intervention combining 30 minutes of paired brain and peripheral nerve stimulation prior to 60 minutes of physical exercise for 20 sessions improved both neurophysiological and functional outcomes in people with chronic SCI at a range of levels and severities (Jo et al. 2023). No studies have conclusively assessed a longitudinal course of multimodal therapy comparing parallel groups receiving stimulation before versus during exercise therapy. However, with an eye on clinical implementation, delivering stimulation as a primer before locomotor training is significantly simpler than delivering stimulation concurrently with locomotor training.

Another unresolved question regarding transspinal stimulation concerns in what *body position* to deliver stimulation as a primer for locomotor training? As opposed to the supine position, delivering stimulation in the upright standing position concurrently engages neural circuits involved in sensing weight bearing and adjusting leg muscle tone accordingly. Standing results in a low-level soleus background activity, even in a body weight-supported condition (Baudry et al. 2014; Tokuno et al. 2009). Soleus background activity increases propriospinal circuit activation, results in more consistent H-reflex latencies, and reduces the level of soleus H-reflex rate-dependent depression (Brangaccio et al. 2025; Burke et al. 1989; Stein et al. 2007). Chronic SCI may reduce soleus H-reflex rate-dependent stimulation. Therefore, it remains unclear whether the background activity induced by supported standing would facilitate or interfere with the benefits of tonic transspinal stimulation and subsequent locomotor training.

As such, we undertook this novel study to directly compare the effects of body posture during transspinal stimulation when used as a ‘primer’ for task-specific walking exercise in a longitudinal study in people with chronic motor-incomplete SCI. All participants underwent 40 sessions (30 minutes each) of weight-supported locomotor training. Participants were randomly assigned to three different forms of priming locomotor training with transspinal stimulation (30 minutes each) over the thoracolumbar enlargement of the spinal cord: active (30 Hz) stimulation in the supine position, active stimulation in the standing position, or sham stimulation in the standing position. Before and after the 40-session protocol, we measured the effects of these three interventions on the strength of homosynaptic depression, reciprocal inhibition, presynaptic inhibition, and recruitment order and excitability threshold of muscle afferents and alpha motoneurons.

## 2 Material and methods

### 2.1 Participants

We recruited adults aged 18-70 years old with motor incomplete (American Spinal Injury Association Impairment Scale (AIS) SCI at least 6 months prior to enrollment, with a lesion level at or rostral to the T10 neurological level. Study participation for each subject was approximately 2.5 months. The inclusion and exclusion criteria are described in detail in the published clinical trial protocol (Skiadopoulos et al. 2023).

All participants signed an informed consent form, which was approved by the Institutional Review Board (IRB) of the City University of New York (CUNY IRB 2019-0806) and James J. Peters Veterans Affairs Medical Center (JJPVAMC) (IRB 01919). Eight participants completed the training at the Klab4Recovery (Knikou) SCI Research Program at CUNY, and 6 participants completed training at JJPVAMC. Neurophysiological assessments were performed at the Klab4Recovery. The work was carried out in accordance with the World Medical Association Declaration of Helsinki. Trial registration: ClinicalTrials.gov: NCT04807764; Registered on March 19, 2021, https://clinicaltrials.gov/ct2/show/NCT04807764.

### 2.2 Intervention: transspinal stimulation before locomotor training within the same session

Each participant received an average of 40 sessions of 30 Hz transspinal stimulation and locomotor training with the Lokomat 6 Pro®. Transspinal stimulation was delivered with a current stimulator (charge-balanced, symmetric, biphasic rectangular pulses of a 1-ms width per phase; DS8R, Digitimer Ltd., UK) based on each participant’s comfort at 1.2 multiples of paresthesia threshold for 30 min per session (Table 1).

**Table 1.** Demographics and injury characteristics of participants with chronic spinal cord injury (SCI).

| Subject ID | Gender | Age range | Vertebra Level of Injury | AIS<sup>1</sup> Scale | Time post injury (yr) | Cause of Injury | Neurotropic Medications | # of sessions attended |
| --- | --- | --- | --- | --- | --- | --- | --- | --- |
| ACTIVE-STANDING |  |  |  |  |  |  |  |  |
| NIH007 | F | 41-45 | C1 | D | 2 | MVA | Baclofen 10mg 5x/day, Cyclobenzaprine 15mg/day<br>Dentrolene 50mg 3x/day<br>Duloxetine 20mg 2x/day | 41 |
| NIH008 | F | 25-30 | T4 | D | 10 | Scoliosis repair surgery | None | 41 |
| NIH009 | M | 60-70 | C3 | D | 3 | Fall in bathroom | Baclofen 35 mg/day, gabapentin 300mg 3x/day, pregabalin 150mg 3x/day, diazepam 0.5 mg 3-4x/day, | 41 |
| NIH011 | M | 40-45 | T5 | D | 2 | Fall from roof | Baclofen 5mg 5x/day, dalfampridine 10mg 2x/day | 43 |
| SHAM-STANDING |  |  |  |  |  |  |  |  |
| NIH001 | M | 60-65 | C2 | B | 11 | Swimming | Baclofen 20mg 1x/day | 40 |
| NIH002 | F | 60-65 | C4 | D | 10 | Swimming | Baclofen 20 mg 2x/day, cyclobenzaprine 5mg 3x/day, oxybutynin 10 mg/day, amitriptyline 10 mg/day | 40 |
| NIH003 | M | 50-55 | T3 | D | 12 | Spinal arachnoid cyst | Baclofen 20 mg 4x/day; testosterone 40.5mg 2x/week | 40 |
| NIH010 | M | 20-25 | T10 | B | 3 | Ski accident | Baclofen 10mg 3x/day, gabapentin 300 mg 2x/day | 42 |
| NIH014 | M | 25-30 | T8 | D | 15 | AVM<sup>4</sup> rupture | Methenamine hippurate 1g 2x/day | 41 |
| ACTIVE-SUPINE |  |  |  |  |  |  |  |  |
| NIH004 | F | 30-35 | C3 | D | 17 | MVA<sup>3</sup> | Bupropion 300 mg 1x/day | 40 |
| NIH005 | M | 45-50 | T8 | D | 22 | Gunshot | Oxycodone 10mg, gabapentin 300mg | 34 |
| NIH006 | F | 25-30 | T2 | B | 8 | Gunshot | Vibegron 75mg 1x/day | 36 |
| NIH012 | M | 55-60 | T3 | D | 1 | Transverse myelitis | Baclofen 25mg 3x/day, Tizanadine 4mg 3x/day, Oxybutinin 10 mg 1x/day | 41 |
| NIH013 | M | 20-25 | C4 | D | 0.5 | Spinal stroke | Myrbetriq 15mg 1x/day, Atorvastatin 20mg 1x/day<br>Cialis 2.5-5mg as needed | 36 |

<sup>1</sup> American Spinal Injury Association Impairment Scale
<sup>2</sup> All groups received 30-minutes of locomotor training with the Lokomat 6 Pro after 30 minutes of active or sham transspinal stimulation during supine or standing.
<sup>3</sup> Motor vehicle accident.
<sup>4</sup> Arteriovenous malformation.

Transspinal stimulation was delivered during standing based on the importance of upright posture regulation in locomotor control (Deliagina et al. 2012; Milosevic et al. 2015, 2017; Lemay et al. 2015), which is greatly affected after SCI. During standing, the soleus H-reflex amplitude is directly related to postural instability and dynamic balance (Kawaish and Domen 2016) and is susceptible to cortical control (Papegaaij et al. 2016; Soto et al. 2006). Specifically, upright balance control requires increased soleus motor evoked potentials and decreased soleus H-reflexes (Baudry et al. 2014; Tokuno et al. 2009). We compared active transspinal stimulation delivered during supported standing versus supine to distinguish whether soleus background activity during tonic transspinal stimulation facilitates or interferes with subsequent locomotor training benefits compared to the supine position. We also compared active versus sham transspinal stimulation delivered during standing to differentiate the additive effects of transspinal stimulation when added to stand training.

For all participants, a single reusable self-adhered cathode electrode (10.2 × 5.1 cm^2^, Uni-Patch, Massachusetts, USA) was placed at midline overlying the vertebrae equally between the left and right paravertebral sides covering from Thoracic 10 to Lumbar 1-2 vertebral levels. The anode electrode was a pair of interconnected electrodes (same type as the cathode) placed on each iliac crest (Knikou et al. 2015b). The Thoracic 10 spinous process was identified via palpation and based on anatomical landmarks (end of rib cage, Thoracic 1 vertebra). Consistent cathodal electrode position across the intervention sessions was ensured by carefully recording the position of the electrode relative to anatomical landmarks. For participants who received transspinal stimulation while lying supine, hips and knees were placed in slight flexion and stabilized by holsters and towels to avoid external limb rotations as needed. Active transspinal stimulation at 30 Hz delivered during standing was administered under body weight support (BWS) in the Lokomat 6 Pro ®. BWS during standing was adjusted to minimize knee buckling and was decreased over the training sessions to achieve full loading. Sham transspinal stimulation during standing consisted of current delivery at above paresthesia threshold for 1 min, ramped slowly to 0 mA intensities that remained for 28 min, followed by ramping back to above paresthesia threshold for the final minute. Over the training course, we used the same clinical algorithm to adjust the BWS, leg guidance force (LGF), and treadmill speed for locomotor training as we have used in our previous clinical trial (Knikou 2013). The tension of the ankle straps, BWS and LGF were adjusted to achieve absence of toe and foot dragging during assisted stepping. Of note, one participant (NIH004) was able to perform treadmill training without robotic assistance. All participants tolerated the intervention well. Several participants had mild skin abrasions during the training protocol, which is expected in exoskeletal-assisted walking protocols. All adverse events were resolved. The blood pressure of all participants was monitored during the intervention and no changes were noted.

### 2.3 Surface EMG

For neurophysiological assessments, the skin was dry shaved, abraded, and cleaned with alcohol. Differential bipolar surface electrodes with fixed inter-electrode distance of 2 cm (Motion Lab System EMG preamplifier) were secured with Tegaderm transparent film (3M Healthcare, St. Paul, MN, USA). We recorded EMG signals from the soleus (SOL) or tibialis anterior (TA) muscles while standing or sitting. All EMG signals were sampled at 2,000 Hz with an EMG unit (MA300 DTU, Motion Lab Systems Inc., Baton Rouge, LA, USA), and acquired using either a 1401 POWER mkII analog-to-digital interface running Spike 2 (Cambridge Electronics Design Ltd., England, UK) or a 16-bit data acquisition card (NI-PCI-6225, National Instruments, Austin, TX) running customized LABVIEW software.

### 2.4 Neurophysiological biomarkers before and after treatment

The neurophysiological tests described below were performed 1 or 2 days after completion of all training sessions. The soleus H-reflex was used as a probe of neuroplasticity and neurorecovery (Knikou 2008). The optimal stimulation site for the soleus H-reflex was established with subjects seated, and corresponded to the site that an H-reflex could be evoked at low stimulation intensities without an M-wave being present, and at increasing stimulation intensities the shape of the M-wave and H-reflex were similar. The optimal stimulation site was established via a custom-made monopolar stainless-steel hand-held electrode (cathode) that was replaced by a permanent pre-gelled disposable electrode (SureTrace, Conmed, NY) maintained under pressure via a custom-made pad and athletic pre-wrap tape.

#### 2.4.1 Homosynaptic depression

Soleus H-reflexes following posterior tibial nerve stimulation with a 1-ms monophasic pulse were recorded with subjects seated and stimuli delivered every 1s (1.0 Hz), 3s (0.33 Hz), 5s (0.2 Hz), 8s (0.125 Hz), and 10s (0.1 Hz). Homosynaptic depression is greatest at 1s and fully recovers at 8 or 10s (Crone and Nielsen 1989). Homosynaptic depression was not recorded during standing and stepping because it is abolished by strong spinal inhibitory circuits and muscle contraction (Burke et al. 1989; Stein et al. 2007). Soleus H-reflexes were evoked and recorded on the ascending part of the recruitment curve with amplitude ranging from 20-35 % of the maximal M-wave, randomly across different frequencies with 15 H-reflexes recorded at each frequency.

#### 2.4.2 Reciprocal and presynaptic inhibition

To assess restoration of reciprocal Ia inhibition and presynaptic inhibition, soleus H-reflexes were recorded following common peroneal nerve (CPN) stimulation at short and medium conditioning-test (C-T) intervals. In seated relaxed subjects, ipsilateral CPN stimulation was delivered with a bipolar stainless-steel electrode placed distal to the head of the fibula to determine the most optimal stimulation site. This site was optimized such that the TA motor threshold was lower than that of the peroneal muscles, and at increased stimulation intensities the peroneus longus muscle was inactive (Knikou 2008). Reciprocal Ia inhibition was assessed with a conditioning single pulse of 1-ms in duration, generated by a constant current stimulator (DS7A, Digitimer, UK), delivered to the CPN at the C-T intervals of 0, 1, 2, 3, and 4 ms. Presynaptic inhibition was assessed with a conditioning pulse train of 4 pulses with 9 ms duration, generated by a constant current stimulator (DS7A, Digitimer, UK) and delivered to the CPN at the C-T intervals of 20, 60, or 100 ms. These C-T intervals were selected because reciprocal Ia inhibition has a short latency between the activation of the agonist and inhibition of the antagonist in humans (Crone et al. 1987), whereas the reflex inhibition produced at the intermediate C-T intervals is predominantly presynaptic (Iles1996). For all cases, constancy of conditioning stimulation was ensured by the presence of a stable, small amplitude TA M-wave, which was monitored during the experiment using a Digital oscilloscope. The stimulus to the CPN was delivered at 1.27 ± 0.12 (21.8 ± 10.1 mA) and 1.4 ± 0.28 (21.8 ± 10.1 mA) TA motor threshold before and after treatment, respectively. Control and conditioned soleus H-reflexes were randomly recorded across the C-T intervals tested, and 15 H-reflexes were recorded at each C-T interval.

#### 2.4.3 Alpha motoneuron excitability

To minimize presence of ankle clonus and spasms upon repetitive stimulation of Ia afferents with participants seated, the soleus H-reflex and M-wave recruitment curves were assembled with subjects standing at a BWS (58 ± 25 %) that prevented knee buckling. A monophasic stimulation pulse was delivered randomly at a wide range of intensities. Recordings were taken at intensities from below Ia afferent threshold until M-waves reached maximal amplitudes. At least 100 pulses were delivered randomly at different stimulation intensities to assemble the H-reflex and M-wave recruitment curves.

### 2.5 Data analysis

For homosynaptic depression, soleus H-reflexes recorded at different stimulation frequencies (0.125, 0.2, 0.33, and 1.0 Hz) were measured as the area under the full-wave rectified waveform and were normalized to the mean amplitude of the homonymous H-reflex evoked at 0.1 Hz. For reciprocal and presynaptic inhibition, soleus H-reflexes conditioned by antagonistic CPN stimulation at short (0, 1, 2, 3, and 4 ms) and medium (20, 60, and 100 ms) C-T intervals were normalized to the mean amplitude of the control H-reflex.

Soleus M-waves and H-reflexes recorded at varying increasing stimulation intensities (recruitment input-output curve) were measured as peak-to-peak of the non-rectified waveform and were normalized to the associated maximal M-wave to counteract differences of muscle geometry across participants. Normalized responses were plotted against the non-normalized stimulation intensities (Smith et al. 2015; Klimstra and Zehr 2008). A Boltzmann sigmoid function (Eq. 1) was fitted to the data to establish the stimulus intensity corresponding to 50 % of maximal H-reflex (S50-Hmax) or 50 % of maximal M-wave (S50-Mmax), the predicted maximal values for the H-reflex and or M-wave, and the slope parameter of the function (m). From the m and S50-Hmax or S50-Mmax, we estimated the slope, and stimulation intensities corresponding to the H-reflex and M-wave thresholds and maximal amplitudes based on equations 2, 3, and 4. These parameters were grouped and averaged based on the study group and time of testing.

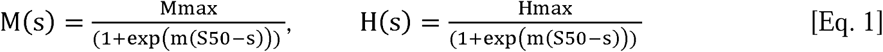

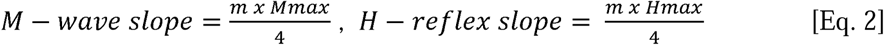

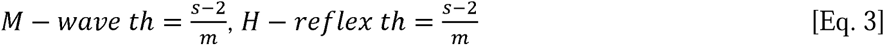

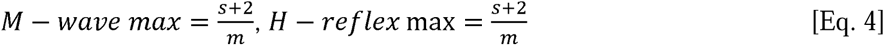

The homonymous predicted S50-Mmax was used to normalize the stimulation intensities and group the M-waves across participants. Because the S50-Hmax after treatment was decreased, the S50-Hmax observed at baseline was used to normalize the stimulation intensities for the soleus H-reflexes recorded before and after intervention (Knikou and Murray 2019). Averages of normalized M-waves and H-reflexes were calculated across multiples of the S50-Mmax or S50-Hmax in steps of 0.05 up to 2.0 and in steps of 1.0 thereafter. This was done separately for each soleus H-reflex recruitment curve. Normalized M-waves and soleus H-reflexes were grouped across subjects based on multiples of S50-Mmax or S50-Hmax, study group, and time of testing.

For each participant, we calculated the average minimum BWS, minimum LGF and maximum speed reached within each block of 5 sessions was calculated. We also calculated the percentage of change in these parameters at the final training block (sessions 36-40) versus the first training block (sessions 1-5).

Outcome measures were analyzed using repeated-measures analysis of variance (rmANOVA) with factors (levels) of group (active-supine, active-standing, sham-standing), time (before or after intervention), and condition (C-T interval or stimulation frequency or multiples of stimulation intensities). A 2-way repeated measures analysis of variance (rmANOVA) or a 3-way rmANOVA was performed to establish significant differences across time within a group or between groups. When statistically significant differences were found, Holm-Sidak pairwise multiple comparisons were performed. Results are presented as mean values and SD. For all statistical tests, the effects were considered significant when *p* < 0.05.

## 3 Results

### 3.1 Participant demographics

Fourteen people with chronic SCI were enrolled in the study. Three participants had neurological deficit grade B (sensory incomplete and motor complete), and 11 had grade D (motor function is preserved, and at least half of key muscle functions below the neurological level of injury have a muscle grade greater than 3 on the International Standards for the Neurological Classification of SCI) (Table 1). The vertebral injury level of SCI ranged from Cervical 2 to Thoracic 10. Despite randomized assignment, there were several differences among groups at baseline (Table 1). The active-standing group (n=4) included two males, two females; all four participants with AIS grade D injury; and two with cervical, two with thoracic SCI. The sham-standing group (n=5) included four males, one female; three participants with AIS grade D and two with AIS grade B injury; and two with cervical, three with thoracic SCI. The active-supine group (n=5) included three males, two females; four with AIS grade D and one with AIS grade B injury; two with cervical, three with thoracic SCI. Time post injury ranged from 6 months to 22 years.

### 3.2 Changes in Homosynaptic Depression

Figure 1A depicts an example of the effects on homosynaptic depression in one participant (NIH011) before and after completing the active transspinal stimulation during standing protocol. Non-rectified raw single soleus H-reflexes evoked every 10s (0.1 Hz) and 1s (1.0 Hz) are shown. At 1.0 Hz, the soleus H-reflex before and after treatment was 28.43 % and 13.54 % of the H-reflex evoked at 0.1 Hz, respectively. The decrease in H-reflex amplitude at 1.0 Hz suggests that the intervention potentiated homosynaptic depression after treatment in this participant.

**Figure 1.**
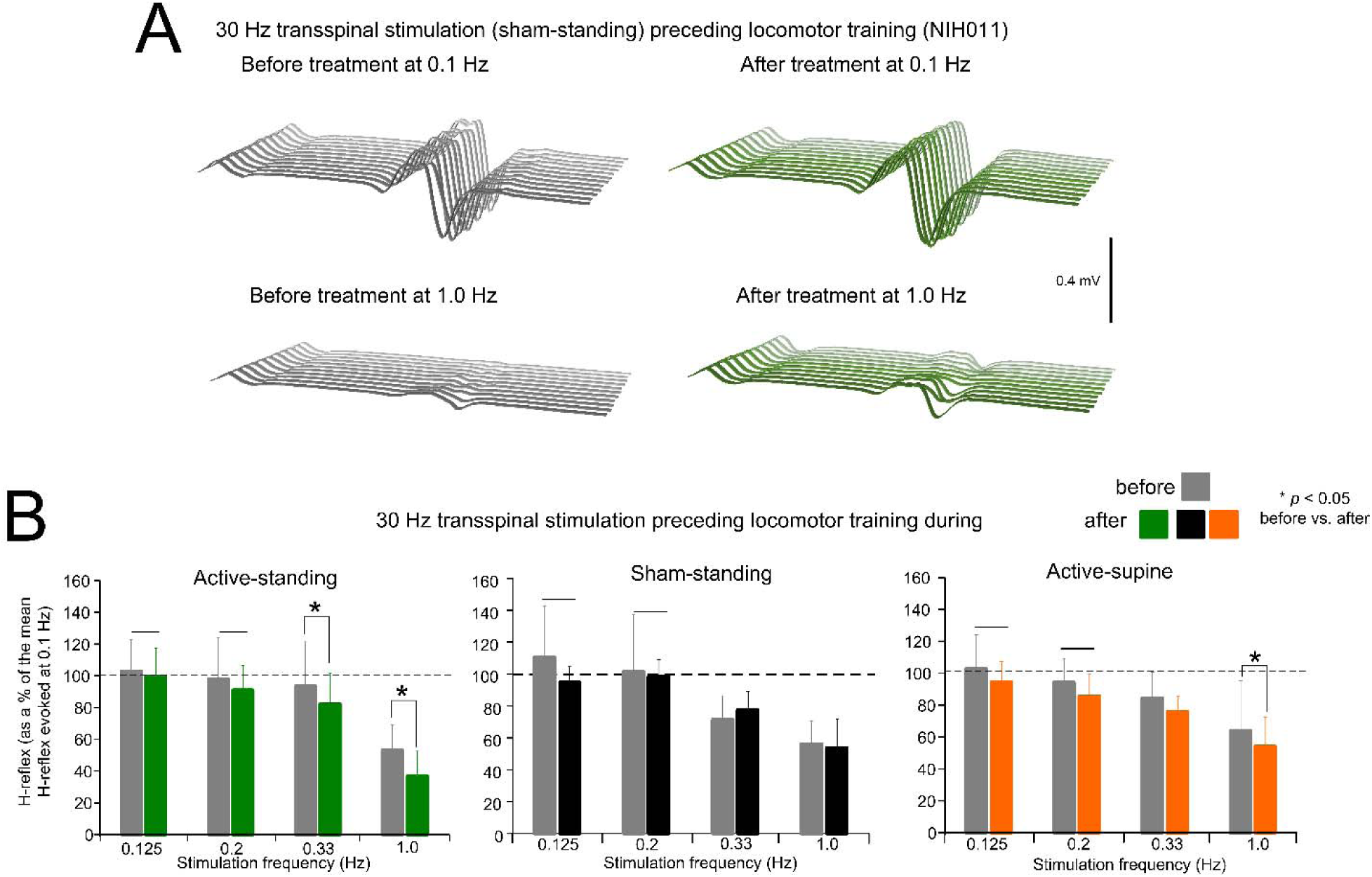
Changes in homosynaptic depression. (**A).** Representative examples of non-rectified raw waveforms of soleus H reflexes recorded at 0.1 and 1.0 Hz before (grey) and after (green) 30 Hz transspinal stimulation preceding locomotor training during standing. All soleus H reflexes are from subject NIH011 who had an AIS D and received a total of 43 training sessions (weekdays). The soleus H-reflex before and after treatment at 1.0 Hz was 28.43 % and 13.54 % of the soleus H-reflex evoked at 0.1 Hz respectively. (**B).** Mean amplitude of the soleus H reflexes evoked at 0.125, 0.2, 0.33, and 1.0 Hz is indicated before and after training for each study group. On the abscissa the stimulation frequency is indicated while the ordinate indicates the amplitude of the soleus H-reflexes as a percentage of the soleus H-reflex evoked at 0.1 Hz. * *p* < 0.05, statistically significant changes of the H reflexes recorded after training compared with those recorded before training at a given stimulation frequency. Horizontal bar (−) indicates the soleus H-reflexes at 0.125 and 0.2 Hz that were significantly different from those recorded at 1.0 Hz before and after treatment. Error bars denote the SD.

The mean amplitude of the soleus H-reflex across participants recorded at 1.0, 0.33, 0.2, and 0.125 Hz as a percentage of the mean amplitude of the H-reflex recorded at 0.1 Hz before and after treatment is indicated in Figure 1B. rmANOVA at 3 (study group) x 2 (time) x 4 (stimulation frequency) levels showed that the soleus H-reflexes varied significantly as a function of time (F_1,167_ = 9.07, *p* = 0.003), and across stimulation frequencies (F_3,167_ = 45.85, *p* < 0.001), but not as a function of the study group (F_2,167_ = 0.02, *p* = 0.97). Holm-Sidak pairwise multiples comparisons showed no statistically significant differences between study groups at the before (*p* > 0.05) or after (*p* < 0.05) treatment time points. An effect of time was apparent within the active-supine and active-standing groups. Specifically, the soleus H-reflexes at 0.33 and 1.0 Hz before and after treatment were significantly different in the active-standing and active-supine groups.

### 3.3 Changes in Presynaptic Inhibition

Figure 2A depicts an example of the effects on presynaptic inhibition in one participant (NIH03) before and after treatment. Non-rectified raw single soleus H-reflex sweeps recorded at the C-T interval of 60 ms were similar or even larger to control reflex values before treatment, while a significant decrease in reflex amplitude was present, suggesting the return of presynaptic inhibition after treatment in this participant.

**Figure 2.**
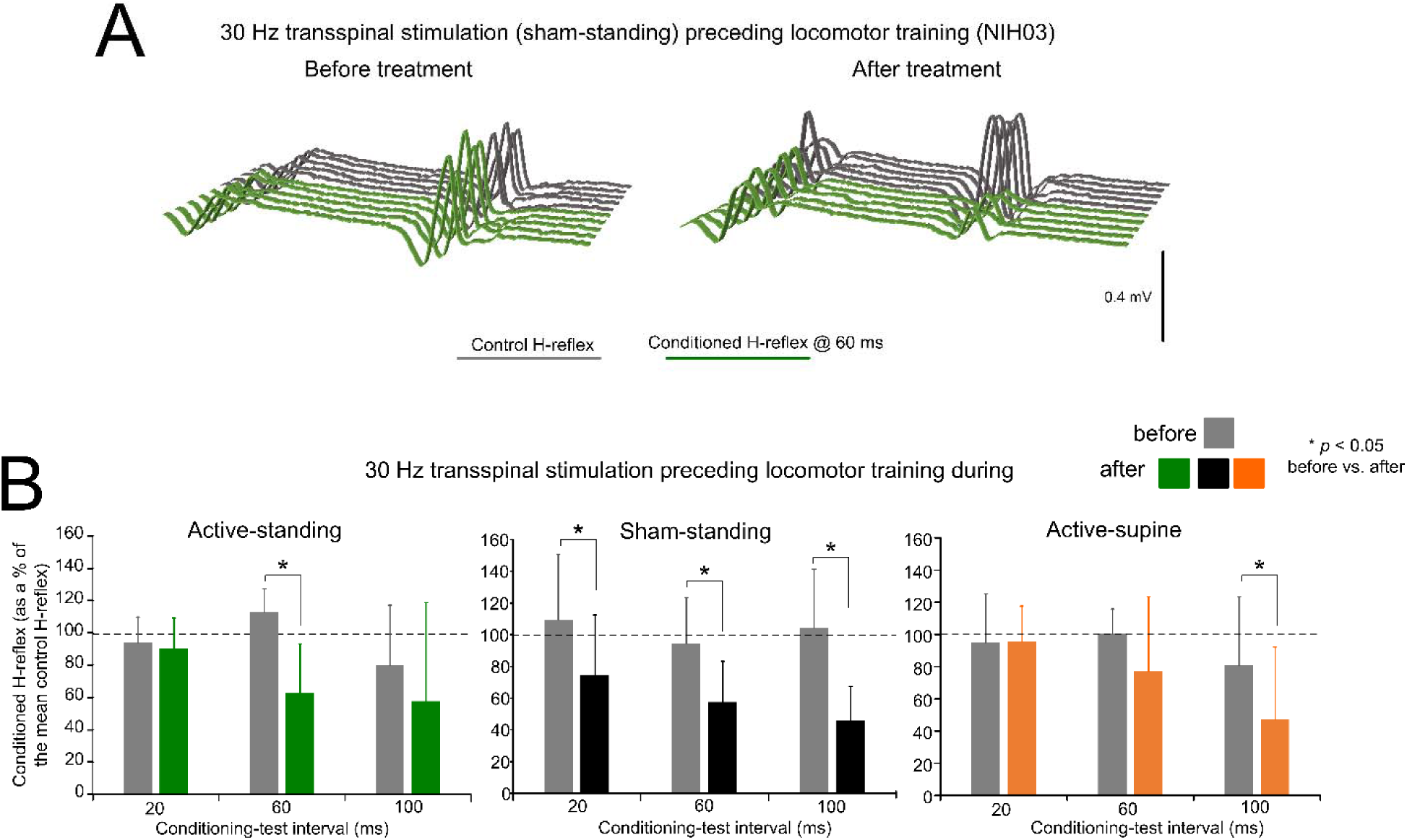
Changes in presynaptic inhibition. (**A).** Non rectified raw sweeps of the soleus H reflexes recorded at the 60 ms conditioning-test interval before and after locomotor training preceded by transspinal stimulation. Control and conditioned H-reflexes are shown for one subject, subject NIH003, who was enrolled in the sham standing study group. The conditioned H-reflex in this subject before and after treatment was 127.8 % and 66.1 % of control reflex values at the C-T interval of 60 ms supporting for profound return of natural presynaptic inhibition of Ia afferents. (**B).** Mean amplitude of the soleus H reflexes conditioned by common peroneal nerve stimulation is indicated before and after treatment grouped for each study group. On the abscissa, the conditioning-test interval is indicated while the ordinate indicates the amplitude of the conditioned soleus H-reflexes as a percentage of the control soleus H-reflex. * *p* < 0.05, statistically significant changes of the conditioned H reflexes recorded before and after treatment. Error bars denote the SD.

The mean amplitude of the conditioned soleus H-reflex by CPN stimulation at the C-T intervals of 20, 60, and 100 ms across participants as a percentage of the homonymous control H-reflex before and after treatment is indicated in Figure 2B. Within the active-standing group, the conditioned soleus H-reflexes were statistically significantly different as a function of time at the C-T interval of 60 ms (F_1,18_ = 3.34, *p* = 0.034). A similar result was found within the sham-standing group, in which the conditioned H-reflexes at the 60 and 100 ms C-T interval were significantly reduced after treatment compared to before treatment (t = 2.59, *p* = 0.015). Last, within the active-supine study group, the conditioned H-reflex was also significantly different after treatment (time: F_1,24_ = 2.09, *p* = 0.04). rmANOVA at 3 (study group) x 2 (time) x 3 (C-T interval) levels showed that the conditioned soleus H-reflex varied significantly as a function of time (F_1,84_ = 14.97, *p* < 0.001) and across C-T intervals (F_2,84_ = 4.33, *p* = 0.016), but not as a function of the study group (F_2,84_ = 1.15, *p* = 0.32). Holm-Sidak pairwise multiples comparisons showed significant differences across time within the active-standing (t = 3.1, *p* = 0.002), active-supine (t = 2.09, *p* = 0.039), and sham-standing (t = 2.84, *p* = 0.006) groups.

### 3.4 Changes in burst EMG activity

CPN conditioning stimulation with either a single monophasic 1 ms pulse or a pulse train of 9 ms in duration prior to tibial nerve stimulation produced rhythmic alternating burst activities in both SOL and TA muscles in some participants. In the examples shown in Figure 3, rhythmic alternating activity in the TA and SOL in subject NIH04 lasted up to 3 seconds before treatment, while after treatment (active supine), the TA rhythmic activity was shortened in duration and the SOL rhythmic activity was abolished. In subject NIH014, prior to the intervention, CPN stimulation at the C-T interval of 60 ms produced 4 - 5 rhythmic bursts in TA muscle lasting 500 ms and a few alternating bursts in SOL muscle that all were abolished after treatment (sham-standing) (Figure 3). These observations suggest that locomotor training ameliorated neurophysiological aspects of spasticity manifested by a reduction in alternating rhythmic activity of antagonistic muscles (clonus/spasms). However, a systematic investigation is needed to determine the actions of transspinal stimulation on the neuronal networks underlying this pathological motor behavior after SCI.

**Figure 3.**
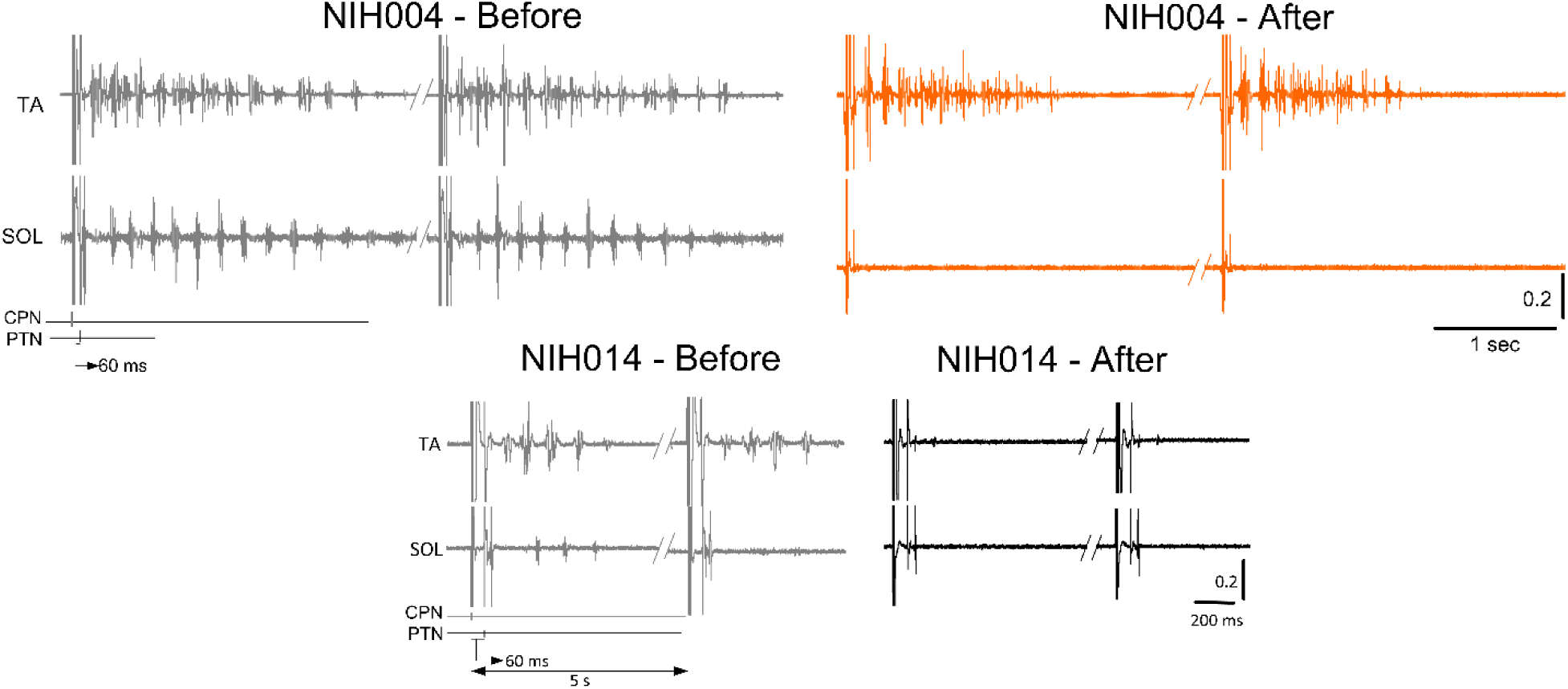
Changes in co-contractions. Raw EMG recordings from the soleus (SOL) and tibialis anterior (TA) muscles upon common peroneal and posterior tibial nerve stimulation at a conditioning-test (C-T) interval of 60 ms. EMG recordings are shown before and after treatment from two representative subjects.

### 3.5 Changes in Reciprocal Inhibition

Figure 4A depicts an example of the effects on reciprocal inhibition in one participant (NIH14) before and after treatment. Representative raw single soleus H-reflex sweeps under control conditions (grey lines) are shown along conditioned reflexes (green lines) recorded at a 2 ms C-T interval. Reciprocal inhibition in subject NIH14 remained unaffected by 41 sessions of sham transspinal stimulation and locomotor training.

**Figure 4.**
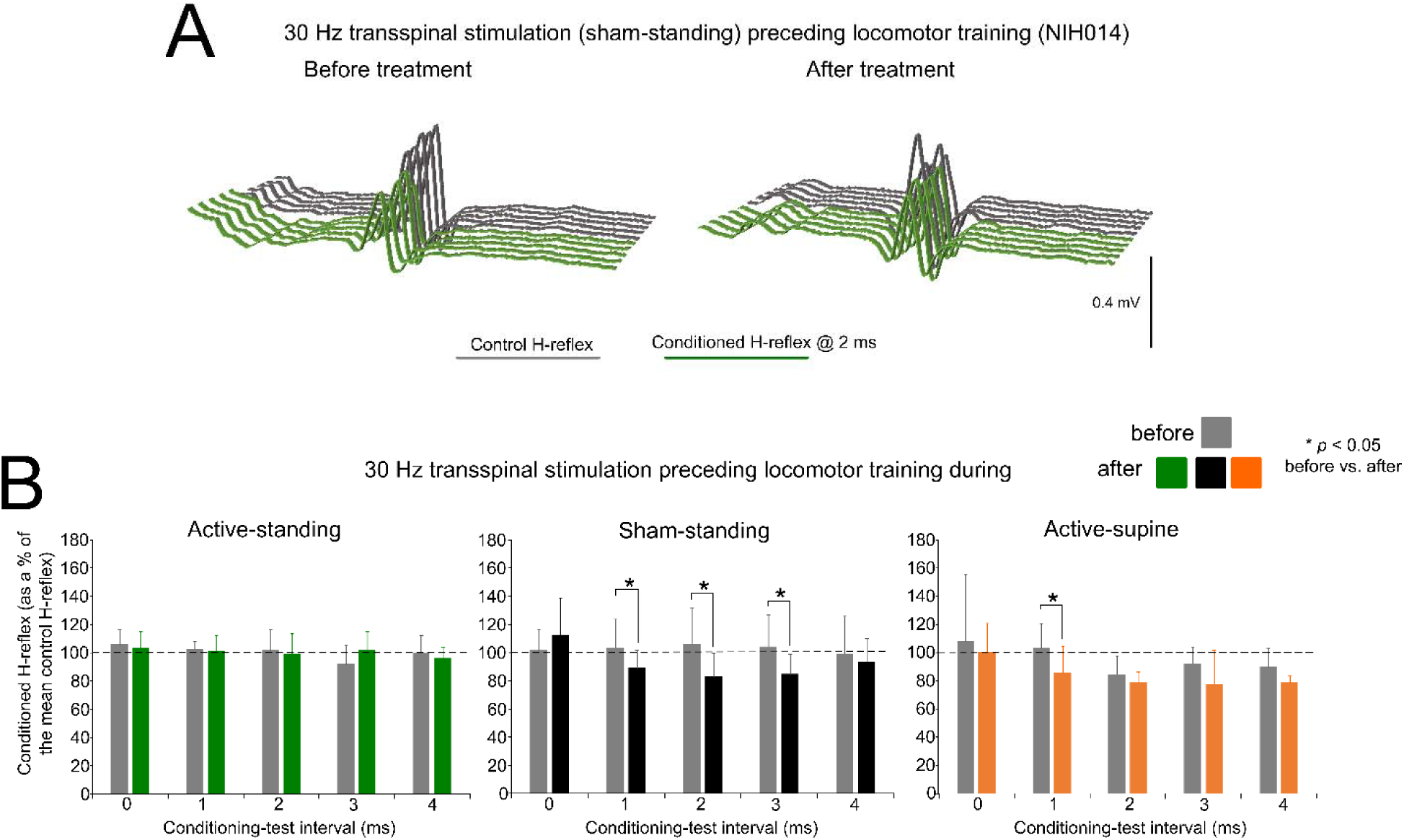
Changes in reciprocal Ia inhibition. (**A).** Non rectified raw sweeps of soleus H reflexes recorded at 2 ms conditioning-test interval before and after priming locomotor training with transspinal stimulation. Control and conditioned H-reflexes are shown for subject NIH014 who was enrolled in the sham-standing group. (**B).** Mean amplitude of the soleus H reflexes conditioned by common peroneal nerve stimulation is indicated before and after treatment grouped per study group. On the abscissa, the conditioning-test (C-T) interval is indicated while the ordinate indicates the amplitude of the conditioned soleus H-reflexes as a percentage of the control soleus H-reflex. * *p* < 0.05, statistically significant changes of the conditioned H reflexes recorded before and after treatment. Error bars denote the SD.

Across participants, the mean amplitude of the conditioned soleus H-reflex by CPN stimulation at short C-T intervals as a percentage of the mean amplitude of the control unconditioned H-reflex before and after 40 sessions of transspinal stimulation and locomotor training are indicated in Figure 4B. Within the active-standing group, the conditioned soleus H-reflexes were not statistically significant different across C-T intervals tested (F_4,26_ = 0.5, *p* = 0.73) or time (F_1,26_ = 0.03, *p* = 0.99). A similar result was also found within the active-supine study group (C-T intervals: F_4,39_ = 1.91, *p* = 0.128; time: F_1,39_ = 3.43, *p* = 0.07). Within the sham-standing group, the conditioned soleus H-reflexes were not significantly different across C-T intervals (F_4,47_ = 0.6, *p* = 0.66) but varied significantly as a function of time (F_1,47_ = 6.54, *p* = 0.014), with H-reflexes at the 2 ms C-T interval being significantly different before and after treatment (t = 27.74, *p* = 0.019).

rmANOVA at 3 (study group) x 2 (time) x 5 (C-T interval) levels showed that the conditioned soleus H-reflex varied significantly as a function of time (F_1,132_ = 6.76, *p* = 0.01), study group (F_2,132_ = 5.17, *p* = 0.007), and C-T intervals (F_4,132_ = 3.44, *p* = 0.01). Holm-Sidak pairwise multiples comparisons showed significant differences between active-standing from active-supine and between sham-standing from active-supine groups. Further, Holm-Sidak pairwise multiples comparisons showed significant differences between time for active-supine (t = 2.41, *p* = 0.017) and sham-standing (t = 2.25, *p* = 0.026) groups.

### 3.6 Changes in Alpha Motoneuron Excitability

Figure 5 shows normalized soleus H-reflexes from all subjects plotted against multiples of S50-Hmax observed at baseline along with the sigmoid fit. In the active-standing group, the soleus H-reflexes were significantly different before and after treatment (F_1,232_ = 10.3, *p* = 0.002), with a significant interaction between time and intensities (F_18,232_ = 1.76, *p* = 0.0031). A similar result was also found for the active-spine group, in which the soleus H-reflexes were significantly different before and after treatment (F_1,242_ = 12.019, *p* < 0.001), with a significant interaction between time and intensities (F_18,242_ = 3.52, *p* < 0.001). Additionally, soleus H-reflexes were significantly different in the sham-standing group before and after treatment (F_1,159_ = 51.59, *p* < 0.001). Two-way rmANOVA at 3 (study group) and 2 (time) levels showed that the soleus H-reflex grouped per multiples of S50-Hmax varied significantly as a function of time (F_1,820_ = 53.59, *p* < 0.001) and among study groups (F_2,820_ = 9.89, *p* < 0.001), while an interaction between study groups and time was evident (F_2,820_ = 5.62, *p* = 0.004). Holm-Sidak pairwise multiples comparisons showed significant differences between active-standing and sham-standing, and between active-standing and active-supine groups. The corresponding soleus M-wave recruitment curves are shown in Figures 5D-F. The soleus M-waves before and after treatment for all three groups were not statistically significant different (active-standing: F_1,259_ = 1.41, *p* = 0.23; active-supine: F_1,246_ = 0.47, *p* = 0.49; sham-standing: F_1,142_ = 0.08, *p* = 0.77).

**Figure 5.**
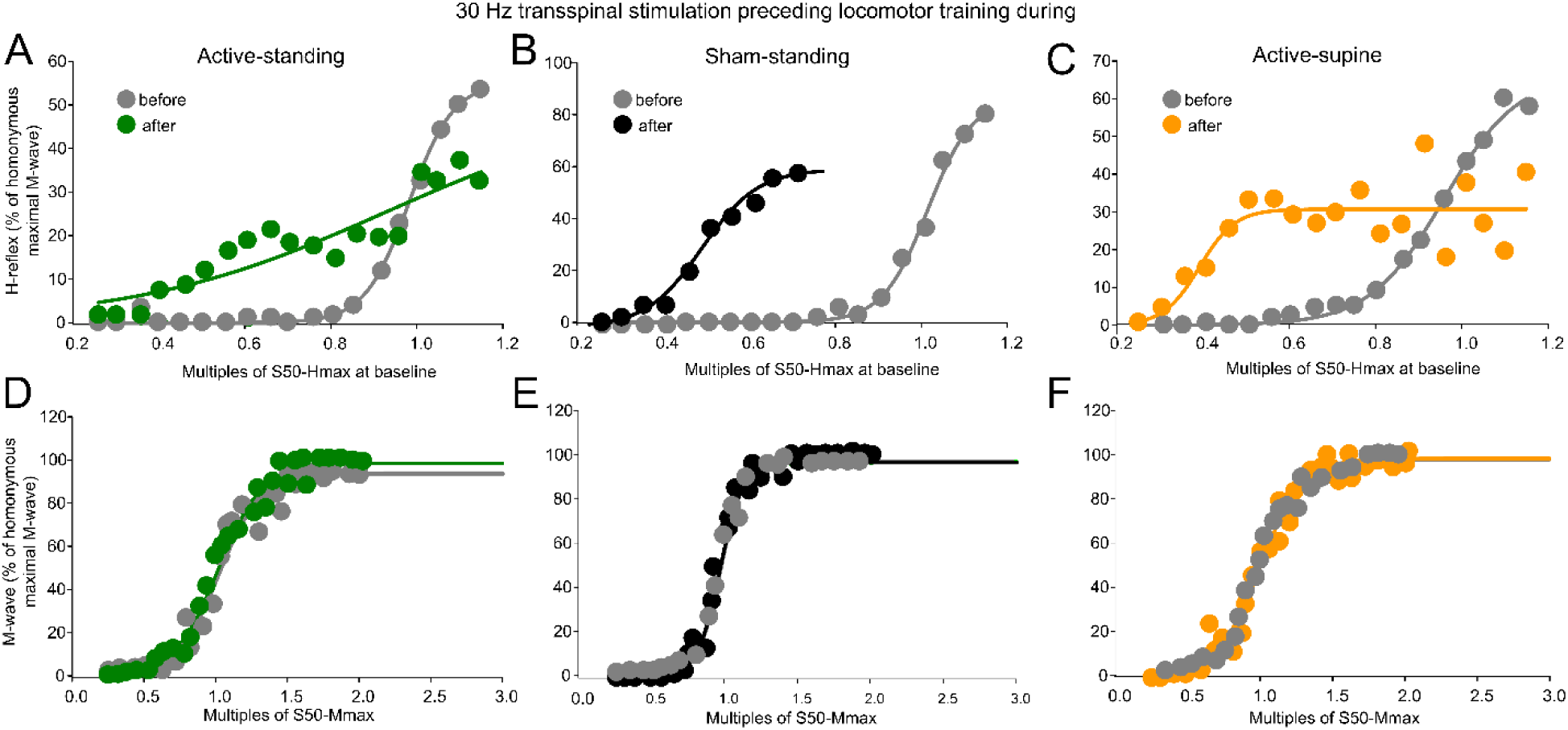
Reflex excitability before and after treatment. (A-C). Overall amplitude of the soleus H-reflexes recorded from below Ia afferent threshold until the H-reflex reached maximal amplitudes is indicated before and after treatment for each study group. On the abscissa, the multiples of stimulation intensities are indicated as normalized values to the S50-Hmax observed at baseline while the ordinate indicates the amplitude of the soleus H-reflexes as a percentage of the homonymous maximal M-wave. **(D-F).** Overall amplitude of the soleus M-waves is indicated before and after treatment for each group. On the abscissa, the multiples of stimulation intensities are indicated as normalized values to the S50-Mmax while the ordinate indicates the amplitude of the soleus M-wave as a percentage of the homonymous maximal M-wave.

Table 2 shows the H-reflex recruitment curve sigmoid fit results. Two-way rmANOVA at 3 (study group) and 2 (time) levels showed that the m function did not vary as a function of time (F_1,42_ = 0.38, *p* = 0.53) or study group (F_2,42_ = 0.49, *p* = 0.61). A similar result was found for the slope (time: F_1,42_ = 4.05, *p* = 0.05; study group: F_2,42_ = 1.42, *p* = 0.25). The stimulation intensities corresponding to S50-Hmax varied significantly as a function of time (F_1,42_ = 7.88, *p* = 0.008) and group (F_2,42_ = 4.52, *p* = 0.01). Similarly, stimulation intensities corresponding to the H-reflex threshold varied significantly as a function of time (F_1,42_ = 7.5, *p* = 0.009) and group (F_2,42_ = 4.58, *p* = 0.01). Stimulation intensities corresponding to the H-reflex maximal amplitude were significantly different between time (F_1,42_ = 7.96, *p* = 0.007) and study group (F_2,42_ = 4.35, *p* = 0.019). For the stimulation intensities corresponding to S50-Hmax, H-threshold and H-max, Holm-Sidak pairwise multiples comparisons showed significant differences between active-standing from active-supine and active-standing from sham-standing groups.

**Table 2.** Sigmoid function predicted parameters for the soleus H-reflex.

|  |  | **R2** | **m** | **S50-Hmax** | **slope** | **Stim at threshold** | **Stim at maximal** |
| --- | --- | --- | --- | --- | --- | --- | --- |
| **Active-standing** | Before | 0.88±0.09 | 2.56±4.56 | 21.13±12.58 | 2.51±1.89 | 18.61±11.53 | 23.63±13.8 |
|  | After | 0.87±0.09 | 1.63±0.8 | 16.01±6.5 | 1.66±1.12 | 14.35±5.51 | 17.67±7.53 |
| **Sham-standing** | Before | 0.89±0.09 | 1.43±0.66 | 15.55±3.83 | 1.86±1.22 | 13.68±3.85 | 17.41±4.19 |
|  | After | 0.89±0.07 | 1.91±0.92 | 8.49±0.62 | 1.24±0.53 | 7.25±0.48 | 9.74±1.04 |
| **Active-supine** | Before | 0.89±0.06 | 1.69±1.67 | 15.04±5.21 | 1.78±0.83 | 13.26±4.56 | 16.82±5.91 |
|  | After | 0.87±0.08 | 3.63±4.01 | 9.28±5.48 | 1.12±0.7 | 8.16±4.9 | 10.41±6.08 |

Sigmoid function parameters were estimated from the sigmoid fitted to the soleus H-reflexes (corresponding to below Ia afferent threshold until the H-reflex reached maximal amplitudes) normalized to the homonymous maximal M-wave and plotted against the stimulation intensities that were normalized to the S50-Hmax observed before treatment. This was performed for each subject separately and values were averaged based on study group. R2 denotes the best fit; m: slope parameter of the function; S50: Stimulus at 50 % of maximal H-reflex. Values are indicated as mean ± SD.

### 3.7 Changes in assisted stepping parameters

Table 3 shows the transspinal stimulation intensities, BWS, LGF, and treadmill speed binned across each set of 5 training sessions. The percentage BWS was significantly different among study groups (F_2,83_ = 9.41, *p* < 0.001) but not among sessions (F_7,83_ = 0.86, *p* = 0.54). Holm-Sidak pairwise multiples comparisons showed significant differences between active-standing and active-supine groups, and between sham-standing and active-supine groups. A similar result was also found for the LGF (study groups: F_2,83_ = 10.45, *p* < 0.001; sessions: F_7,83_ = 0.59, *p* = 0.75), with significant differences found between sham-standing and both active groups. The treadmill speed did vary among study groups (F_2,86_ = 4.21, *p* = 0.018) but not among sessions (F_7,86_ = 0.24, *p* = 0.97), with significant differences found between sham-standing and active-standing groups. The overall percentage of change of the BWS, LGF, and treadmill speed at 36-40 sessions from those observed at 0-5 sessions are presented per study group in Figure 6. The BWS (F_2_ = 9.13, *p* = 0.87), LGF (F_2_ = 3.85, *p* = 0.05), and treadmill speed (H_2_ = 0.32, *p* = 0.85) were not significantly different among groups.

**Figure 6.**
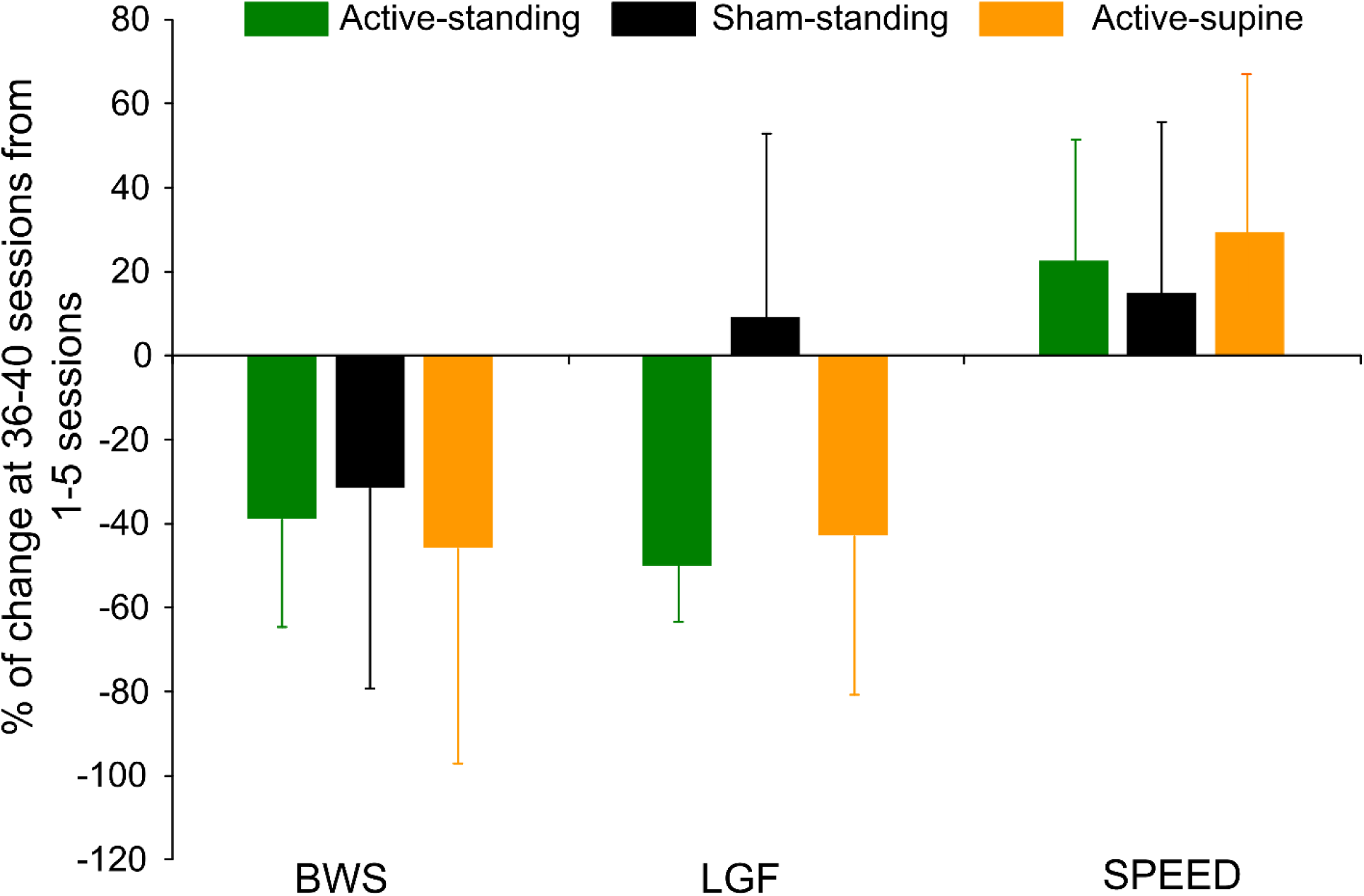
Overall percentage of change of the last block of training sessions (36-40) from the first block of training sessions (1-5) for body weight support (BWS), leg guidance force (LGF), and treadmill speed are indicated for each study group. Improvements in stepping parameters within a group were noted but no statistically significant differences among groups were found. Error bars denote the SD.

**Table 3.** Transspinal stimulation intensities and locomotor training parameters.

| Subject | Sessions | Actual stimulation intensity (mA) | BWS (%) | Speed (m/s) | LGF (%) |
| --- | --- | --- | --- | --- | --- |
| **ACTIVE-STANDING** |  |  |  |  |  |
| NIH007 | 1-5 | 233.2 | 73.4 | 0.28 | 96 |
|  | 6-10 | 358.4 | 61.4 | 0.31 | 79 |
|  | 11-15 | 330.4 | 72.2 | 0.31 | 73 |
|  | 16-20 | 330 | 63.4 | 0.32 | 69 |
|  | 21-25 | 299.2 | 64.2 | 0.29 | 87 |
|  | 26-30 | 332.2 | 73.8 | 0.32 | 80 |
|  | 31-35 | 312 | 52.0 | 0.33 | 55 |
|  | 36-41 | 327.4 | 68.4 | 0.32 | 56.6 |
|  | **% of change (last from 1<sup>st</sup>)** |  | **-6.81** | **14.29** | **-41.04** |
| NIH008 | 1-5 | 99.2 | 31.6 | 0.41 | 86 |
|  | 6-10 | 98.4 | 28.6 | 0.54 | 82 |
|  | 11-15 | 114.6 | 30.8 | 0.47 | 75 |
|  | 16-20 | 114.1 | NR | 0.39 | NR |
|  | 21-25 | 134.4 | NR | 0.39 | NR |
|  | 26-30 | 105.6 | NR | 0.53 | NR |
|  | 31-35 | 91 | 20.5 | 0.58 | 55 |
|  | 36-41 | 95.2 | 14.5 | 0.52 | 48.3 |
|  | **% of change (last from 1<sup>st</sup>)** |  | **-54.11** | **26.83** | **-43.84** |
| NIH009 | 1-5 | 136 | 14.0 | 0.58 | 52 |
|  | 6-10 | 129.1 | 5.0 | 0.56 | 22 |
|  | 11-15 | 116.4 | 5.0 | 0.53 | 20 |
|  | 16-20 | 117.8 | 5.0 | 0.53 | 13 |
|  | 21-25 | 132.2 | 5.0 | 0.52 | 10 |
|  | 26-30 | 142.5 | 5.0 | 0.50 | 11 |
|  | 31-35 | 138.7 | 5.0 | 0.52 | 17 |
|  | 36-42 | 140.9 | 5.0 | 0.52 | 15.7 |
|  | **% of change (last from 1<sup>st</sup>)** |  | **-64.29** | **-10.34** | **-69.81** |
| NIH011 | 1-5 | 236 | 66.6 | 0.34 | 93 |
|  | 6-10 | 221.8 | 59.6 | 0.32 | 73 |
|  | 11-15 | 315.5 | 53.8 | 0.51 | 63 |
|  | 16-20 | 383 | 49.6 | 0.54 | 60 |
|  | 21-25 | 385.5 | 53.4 | 0.42 | 72.5 |
|  | 26-30 | 399.5 | 52.0 | 0.38 | 46.2 |
|  | 31-35 | 330.2 | 50.6 | 0.33 | 49.5 |
|  | 36-43 | 315.3 | 46.0 | 0.54 | 50.6 |
|  | **% of change (last from 1<sup>st</sup>)** |  | **-30.93** | **58.82** | **-45.59** |
| **SHAM-STANDING** |  |  |  |  |  |
| NIH001 | 1-5 |  | 61.3 | 0.57 | 56.2 |
|  | 6-10 |  | 53.0 | 0.66 | 51 |
|  | 11-15 |  | 36.0 | 0.66 | 58 |
|  | 16-20 |  | 32.0 | 0.68 | 79 |
|  | 21-25 |  | 39.0 | 0.62 | 88 |
|  | 26-30 |  | 46.0 | 0.59 | 100 |
|  | 31-35 |  | 50.0 | 0.59 | 100 |
|  | 36-40 |  | 48.0 | 0.56 | 100 |
|  | **% of change (last from 1<sup>st</sup>)** |  | **-21.7** | **-1.75** | **77.94** |
| NIH002 | 1-5 |  | 58.8 | 0.51 | 59 |
|  | 6-10 |  | 47.8 | 0.58 | 61.5 |
|  | 11-15 |  | 38.4 | 0.60 | 65 |
|  | 16-20 |  | 38.0 | 0.60 | 62 |
|  | 21-25 |  | 24.0 | 0.52 | 64 |
|  | 26-30 |  | 16.2 | 0.51 | 64 |
|  | 31-35 |  | 15.8 | 0.52 | 57.5 |
|  | 36-40 |  | 11.4 | 0.48 | 51 |
|  | **% of change (last from 1<sup>st</sup>)** |  | **-80.61** | **-5.88** | **-13.56** |
| NIH003 | 1-5 |  | 32.0 | 0.61 | 84 |
|  | 6-10 |  | 33.0 | 0.66 | 90 |
|  | 11-15 |  | 44.0 | 0.68 | 98 |
|  | 16-20 |  | 45.0 | 0.68 | 100 |
|  | 21-25 |  | 45.0 | 0.70 | 100 |
|  | 26-30 |  | 45.0 | 0.67 | 100 |
|  | 31-35 |  | 45.0 | 0.64 | 100 |
|  | 36-40 |  | 45.0 | 0.63 | 100 |
|  | **% of change (last from 1<sup>st</sup>)** |  | **40.63** | **3.28** | **19.06** |
| NIH010 | 1-5 |  | 76.3 | 0.42 | 100 |
|  | 6-10 |  | 66.6 | 0.41 | 100 |
|  | 11-15 |  | 64.4 | 0.38 | 100 |
|  | 16-20 |  | 58.0 | 0.45 | 100 |
|  | 21-25 |  | 58.0 | 0.43 | 100 |
|  | 26-30 |  | 56.6 | 0.47 | 100 |
|  | 31-35 |  | 48.8 | 0.39 | 100 |
|  | 36-42 |  | 47.7 | 0.38 | 100 |
|  | **% of change (last from 1<sup>st</sup>)** |  | **-27.48** | **-9.52** | **0.0** |
| NIH014 | 1-5 |  | 41.4 | 0.32 | 86 |
|  | 6-10 |  | 67.8 | 0.37 | 96 |
|  | 11-15 |  | 29.4 | 0.47 | 79 |
|  | 16-20 |  | 17.8 | 0.56 | 62 |
|  | 21-25 |  | 20.6 | 0.56 | 73 |
|  | 26-30 |  | 18.6 | 0.63 | 49 |
|  | 31-35 |  | 14.6 | 0.58 | 60 |
|  | 36-41 |  | 13.0 | 0.60 | 53.3 |
|  | **% of change (last from 1<sup>st</sup>)** |  | **-68.6** | **87.5** | **-38.02** |
| **ACTIVE-SUPINE** |  |  |  |  |  |
| NIH004 | 1-5 | 69 | 19.1 | 0.14 | Manual assistance |
|  | 6-10 | 67 | 6.6 | 0.16 |  |
|  | 11-15 | 67 | 4.2 | 0.18 |  |
|  | 16-20 | 71.2 | 3.8 | 0.20 |  |
|  | 21-25 | 72.2 | 3.1 | 0.20 |  |
|  | 26-30 | 69.05 | 1.8 | 0.22 |  |
|  | 31-35 | 69.75 | 2.8 | 0.26 |  |
|  | 36-40 | 70.5 | 1.2 | 0.30 |  |
|  | **% of change (last from 1<sup>st</sup>)** |  | **-93.72** | **114.29** | **N/A** |
| NIH005 | 1-5 | 222.2 | 40.6 | 0.53 | 78 |
|  | 6-10 | 242 | 32.0 | 0.54 | 51 |
|  | 11-15 | 202.5 | 31.4 | 0.56 | 35 |
|  | 16-20 | 215.4 | 29.0 | 0.56 | 30 |
|  | 21-25 | 240.3 | 21.5 | 0.56 | 24 |
|  | 26-30 | 361.2 | 15.6 | 0.56 | 19 |
|  | 31-34 | 384 | 12.8 | 0.56 | 15 |
|  | **% of change (last from 1<sup>st</sup>)** |  | **-68.47** | **5.66** | **-80.77** |
| NIH006 | 1-5 | 114 | 31.0 | 0.42 | 100 |
|  | 6-10 | 114 | 26.3 | 0.42 | 100 |
|  | 11-15 | 88.84 | 39.0 | 0.42 | 100 |
|  | 16-20 | 74.88 | 42.5 | 0.41 | 97 |
|  | 21-25 | 77.52 | 39.0 | 0.37 | 97 |
|  | 26-30 | 78.48 | 41.7 | 0.38 | 100 |
|  | 31-35 | 74.4 | 34.0 | 0.40 | 100 |
|  | 36-41 | 82.4 | 43.1 | 0.36 | 100 |
|  | **% of change (last from 1<sup>st</sup>)** |  | **39.03** | **-14.29** | **0.0** |
| NIH012 | 1-5 | 59.1 | 38.6 | 0.59 | 100 |
|  | 6-10 | 67.1 | 46.0 | 0.74 | 85 |
|  | 11-15 | 64 | 41.4 | 0.68 | 72 |
|  | 16-20 | 61.8 | 33.6 | 0.77 | 66 |
|  | 21-25 | 68.8 | 33.6 | 0.78 | 72 |
|  | 26-30 | 64.2 | 27.3 | 0.87 | 69 |
|  | 31-35 | 55.6 | 23.7 | 0.89 | 73 |
|  | 36-41 | 66.5 | 23.1 | 0.89 | 76.7 |
|  | **% of change (last from 1<sup>st</sup>)** |  | **-40.16** | **50.85** | **-23.3** |
| NIH013 | 1-5 | 150 | 20.0 | 0.63 | 70 |
|  | 6-10 | 158.6 | 14.8 | 0.66 | 49 |
|  | 11-15 | 191.9 | 11.6 | 0.71 | 47 |
|  | 16-20 | 180.1 | 9.2 | 0.71 | 35 |
|  | 21-25 | 182.5 | 7.0 | 0.68 | 27.5 |
|  | 26-30 | 185.5 | 6.4 | 0.62 | 29 |
|  | 31-36 | 167.5 | 6.7 | 0.56 | 22.5 |
|  | **% of change (last from 1<sup>st</sup>)** |  | **-66.5** | **-11.11** | **-67.86** |

For each participant, the average minimum body weight support (BWS), minimum leg guidance force (LGF) and maximum treadmill speed reached within each block of 5 sessions is indicated. The percentage of change in these parameters at the final training block (sessions 36-40) versus the first training block (sessions 1-5) is also indicated.

## 4 Discussion

This is the first comprehensive mapping of the neurophysiological effects of a longitudinal rehabilitation intervention composed of transspinal stimulation administered before locomotor training within the same session in people with any form of central nervous system injury. We report here for the first time that transspinal stimulation administered daily before locomotor training promotes a more physiological state of alpha motoneuron excitability and spinal network function after SCI, coinciding with reduced BWS required to step, increased ability to step at higher treadmill speeds, and less guidance force needed from robotic legs during step training.

The soleus H-reflex is a neurophysiological biomarker for hyperreflexia, spasticity, and spinal circuitry underlying coordination of muscular agonists and antagonists. Homosynaptic depression of the soleus H-reflex was present although at reduced levels in all study groups before treatment, consistent with our prior findings (Knikou and Murray 2019). Transspinal stimulation delivered during standing or supine preceding locomotor training potentiated the homosynaptic depression exerted on Ia afferent terminals (Figure 1B), whereas sham stimulation did not. While homosynaptic depression is exerted at the motoneuron synapse and thus is presynaptic, it is different from the presynaptic inhibition brought up by a conditioning stimulus to a heteronymous nerve that engages activity of spinal inhibitory interneurons. Presynaptic inhibition of Ia afferent terminals was potentiated in all three groups (Figure 2B), while reciprocal Ia inhibition, which is exerted postsynaptically, was potentiated after treatment in the active-supine and sham-standing groups (Figure 4B). It is worth noting that CPN stimulation induced co-contractions of ankle antagonistic muscles were reduced significantly in all groups (Figure 3). These changes may be attributed to strengthening of spinal inhibitory networks while a more physiological balance in the proportion of inhibitory and excitatory synaptic inputs to spinal motoneurons is achieved (Ichiyama et al. 2011). Moreover, potentiation of GABAergic and glycinergic inhibitory neurotransmission/reception, altered intrinsic properties of motoneurons and improvements in synaptic inputs from Ia afferents may each have contributed to these changes (Petruska et al. 2007).

Locomotor training alone lowers hyperpolarized resting membrane potentials of motoneurons, decreases spike trigger threshold levels (membrane potential at which an action potential is triggered), and increases amplitudes of afterhyperpolarization (reflecting a decrease in membrane excitability) (Beaumont et al. 2008; Beaumont and Gardiner 2002, 2003). Moreover, locomotor training alone increases glial cell-derived neurotrophic factor (GDNF) levels in spinalized rats (Côté et al. 2011). In a similar manner, polarizing current passing across the spinal cord modifies the membrane potential of muscle spindle afferents as well as the intrinsic properties and excitability of alpha motoneurons and muscle spindle afferents (Eccles et al. 1962). Multiple sessions of transspinal stimulation alone in spinalized rats prevents potassium-chloride cotransporter isoform 2 (KCC2) membrane downregulation in lumbar motoneurons, that coincided with decreased hyperreflexia and increased low frequency-dependent modulation of the soleus H-reflex (Malloy and Côté 2024).

Changes in motoneuron and Ia afferent excitability were apparent in this study, with the soleus H-reflex recruitment curve shifting to the left (Figure 5), and stimulation intensities corresponding to the H-reflex threshold, 50 and 100 % Hmax, decreasing in all groups (Table 2). Because these changes occurred with stable M-wave recruitment curves, the changes in reflex excitability state and excitability threshold cannot be due to excitation of different groups of afferents and soleus motoneurons by the Ia afferent volleys. The shift to the left of the H-reflex recruitment curves suggests that the threshold for discharge of alpha motoneurons is also decreased probably resulting in a more homogenous recruitment of small motoneurons from the pool and those residing at the subliminal fringe.

Thus, transspinal stimulation and locomotor training can each result in a mixture of modulated afferent fiber, motoneuron, and spinal interneuron excitability. At this point, a logical concern is whether these neurophysiological changes can be attributed to transspinal stimulation alone, locomotor training alone, stand training, or their combination; and whether body posture during stimulation plays a role. In this study, homosynaptic depression was doubled when compared to that observed in our past studies of locomotor training only (Knikou and Mummidisetty 2014) or multiple sessions of transspinal stimulation only (Knikou and Murray 2019) (Figure 1S). Similarly, the percentage of change of the conditioned soleus H-reflex by CPN stimulation at 60 and 100 ms was also doubled when compared to locomotor training only (Knikou and Mummidisetty 2014), while no studies exist reporting on this mechanism following transspinal stimulation only. Reciprocal Ia inhibition, however, did not improve in this study when compared to locomotor training only (Knikou et al. 2015), while evidence for this spinal circuit after transspinal stimulation training only are lacking. Last, a past study of locomotor training only minimally affected the excitability thresholds of Ia afferents (Figure 1S) (Smith et al. 2015). Transspinal stimulation training only increased the stimulation intensities corresponding to H-reflex threshold, 50 and 100 % of Hmax (Knikou and Murray 2019), in opposition to the reduced excitability thresholds we observed here in all groups. These comparisons suggest that the current protocol combining transspinal stimulation as a primer for locomotor training produces more pronounced changes in the function of spinal inhibitory circuits critical for standing and stepping (Malloy et al. 2022) when compared to each intervention administered alone.

Significant differences and similarities in the reorganization of spinal inhibitory circuits were evident across study groups. For example, homosynaptic depression was increased in the active-standing and active-supine groups but not in the sham-standing group, suggesting that transspinal stimulation accounted mostly for this neuronal reorganization since all groups received locomotor training. In contrast, reciprocal Ia inhibition was increased in the sham-standing group but not in the active-standing and active-supine groups. These effects may be related to the SCI grade because reciprocal Ia inhibition improves more in response to locomotor training in people with AIS C compared to AIS D injury grades (Smith et al. 2015). However, reciprocal inhibition was not significantly different at baseline between study groups and thus it is difficult to attribute the effects to a specific factor. Presynaptic inhibition as well as excitation of Ia afferents and orderly recruitment of soleus alpha motoneurons were reorganized in a similar manner in all three study groups, suggesting complicated interactions between different levels of the multimodal intervention we used here.

Regarding the multimodal intervention we employed in this clinical trial, whether delivery of transspinal stimulation is best applied before, during, or after locomotor training sessions remains unresolved (Comino-Suárez et al. 2025). Similarly, whether the strategy of alternating subthreshold with suprathreshold intensities at low frequencies during tonic stimulation (Malloy and Côté, 2024; Malloy et al., 2022; Murray and Knikou, 2019) is superior or equal to continuous 30 Hz is also unknown. Testing these options was outside the scope of the current trial, in which we chose to use a priming strategy of active or sham stimulation before locomotor training, while comparing the effects of varied body posture. No studies have conclusively assessed a longitudinal course of multimodal therapy comparing parallel groups receiving stimulation before versus during exercise therapy. However, with an eye on clinical implementation, delivering stimulation as a primer before locomotor training is significantly simpler than delivering stimulation concurrently with locomotor training.

### 4.1 Limitations

The main limitation of this study was the small number of participants who completed the intervention. Not only did this reduce statistical power, but it also resulted in an imbalance of AIS grades and chronicity of SCI across groups (Table 1). Two of five participants in the sham-standing group had AIS grade B injuries, whereas all four participants in the active-standing group had AIS grade D injuries. Likewise, four of the five participants randomized to sham-standing had injury durations of 10 or more years, whereas three of four participants randomized to active-standing had injury durations of 3 or fewer years. Participants with shorter durations of injury and higher levels of function at baseline tend to respond better to activity-based therapeutic interventions (Jones et al. 2014). Consistent with this trend, it was also evident that the active-standing group had higher spinal neuronal network baseline excitability compared to the other study groups. We thus suggest that the depth of neuronal reorganization due to activity-dependent plasticity (stimulation + exercise) depends on the baseline excitability state of motoneurons.

## 5 Conclusion

This randomized control trial assessed the impact of a 40-session intervention combining 30 minutes of active or sham lumbar transspinal stimulation preceding 30 minutes of assisted locomotor training in people with chronic incomplete spinal cord injury. Participants were randomized into three groups: active stimulation in the supine position prior to locomotor training; active stimulation in the weight-bearing upright position prior to locomotor training; and sham stimulation in the weight-bearing upright position prior to locomotor training. Though limited by small numbers of enrollees, we observed evidence for reorganized spinal circuitry after the intervention – all groups demonstrated partially restored presynaptic inhibition; groups receiving active stimulation demonstrated partially restored homosynaptic depression; and all groups demonstrated decreased soleus H-reflex threshold. These findings indicate the potential for multimodal transspinal stimulation and locomotor therapy to improve spinal cord reflex function and potentially improve clinical recovery after chronic SCI.

## Data Availability

All data produced in the present study are available upon reasonable request to the corresponding author.

## Data availability statement

Data will be made available on reasonable requests. Requests to access these datasets should be directed to Maria Knikou.

## Ethics statement

Ethical approval was granted from the City University of New York and James J. Peters Veterans Affairs Medical Center IRB committees. Written informed consent to participate in this study was provided by all participants before study enrollment.

## Author contributions

Conceptualization: MK; Data curation: ASA, MK; Formal analysis: ASA, MZ, MK; Funding acquisition: MK, NYH; Investigation: ASA, MZ, MK; Methodology: MK; Project administration: MK, NYH; Resources: MK, NYH; Software: MK; Supervision: MK, NYH; Validation: MK; Visualization: MK; Writing-original draft: MK; Writing-review and editing: MK, NYH

## Acknowledgements

We are grateful to all research participants, their families and caregivers for their dedication during the study. We thank the staff of the Klab4Recovery SCI Research Program for their assistance in data acquisition and training, and the staff of the James J. Peters VA Medical Center for their assistance in training. We also thank Dr. Marie-Pascale Côté for reviewing a previous version of this paper.

---

**Figure 1S.**
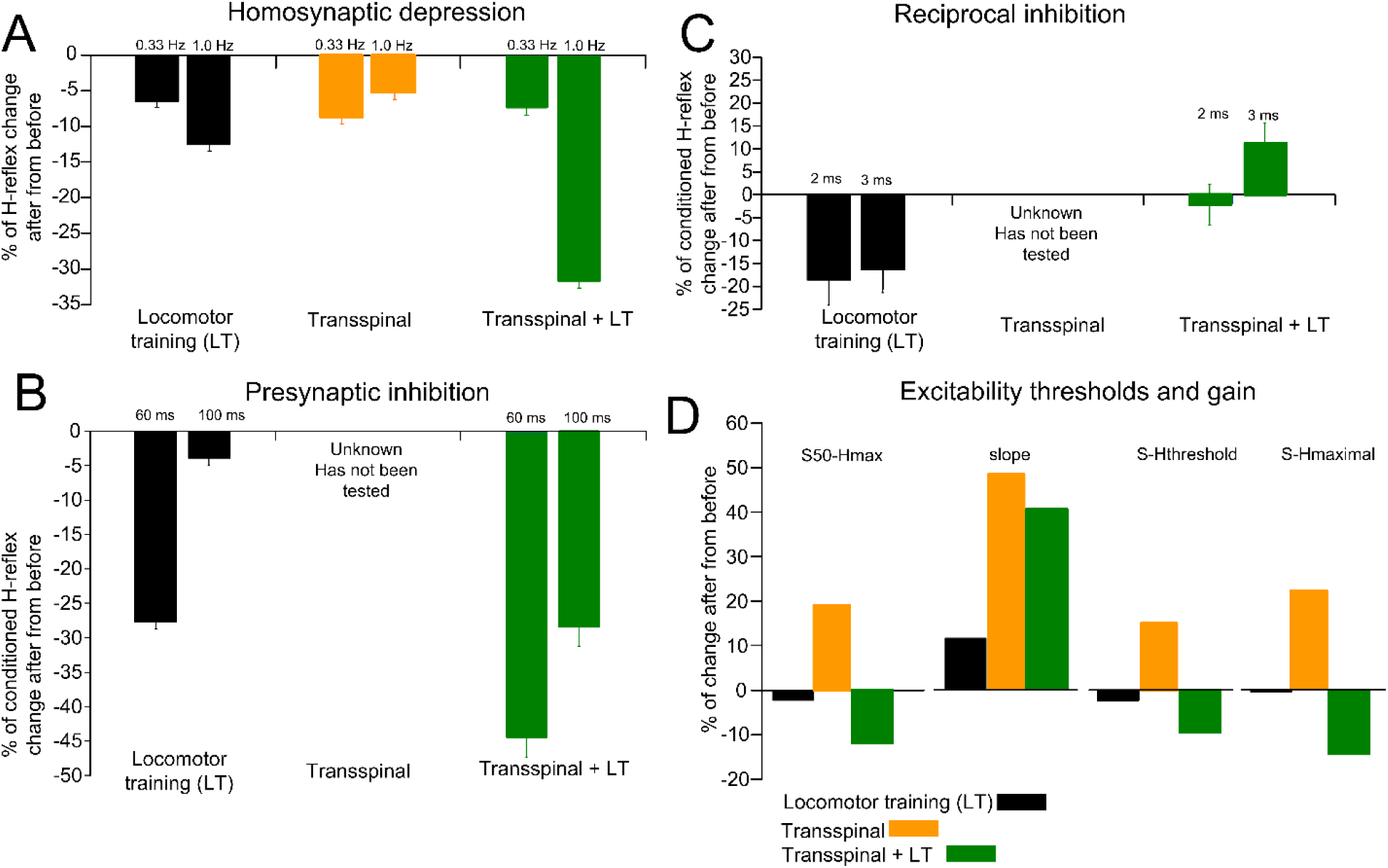
Reorganization of spinal inhibitory circuits and excitability for different interventions. The percentage of changes after intervention from baseline **(A)** low-frequency dependent depression estimated from the soleus H-reflex amplitude evoked at 0.33 and 0.1 Hz, **(B)** presynaptic inhibition estimated by the soleus H-reflex depression by antagonistic peroneal nerve stimulation at 60 and 100 ms C-T intervals, **(C)** reciprocal inhibition estimated by the soleus H-reflex depression by antagonistic peroneal nerve stimulation at 2 and 3 ms C-T intervals, and **(D)** excitability thresholds and gain estimated from the soleus H-reflex recruitment curve are shown for clinical trials that administered locomotor training (LT) alone, transspinal stimulation alone at rest, and transspinal stimulation delivered during standing before LT within the same session (this study). It is apparent that transspinal stimulation + LT nearly doubles the homosynaptic depression and presynaptic inhibition exerted on soleus Ia afferents, while the stimuli corresponding to 50% Hmax, H-threshold, and maximal H-reflex tend to have a similar adaptation between LT and transspinal + LT trials. Only the slope was increased in a similar manner in all three different interventions. Data adopted and re-analyzed from the current study and from Knikou and Mummidisetty 2014, Smith et al. 2015, Knikou et al. 2015a, Knikou and Murray 2019.

